# Household composition, smoking cessation and relapse: results from a prospective longitudinal Australian cohort

**DOI:** 10.1101/2022.01.03.22268695

**Authors:** Karinna Saxby, Andrew Ireland, Peter Ghijben, Rohan Sweeney, Kah-Ling Sia, Esa Chen, Michael Farrell, Hayden McRobbie, Ryan Courtney, Dennis Petrie

**Affiliations:** Centre for Health Economics, Monash Business School, Monash University, Building H, 900 Dandenong Road Caulfield East, Victoria, Australia; National Drug and Alcohol Research Centre, University of New South Wales, R1 Building, 22-32 King St Randwick, New South Wales, Australia

## Abstract

**Aims:** To examine the association between household members and their tobacco smoking behaviour on patterns of smoking cessation and relapse.

**Design and participants:** Data was sourced from 19 waves (years 2001 to 2019) of the nationally representative Household Income and Labour Dynamics in Australia (HILDA) survey, with all household members 15 years or older completing the survey annually. The final sample included, on average, 3,056 ex-smokers and 2,612 smokers per wave.

**Measurements:** Self-reported annual smoking status was used to construct measures of smoking cessation and relapse. Information on household structure and relationships was then used to develop variables describing the presence of household members and their smoking status by relationship to the individual (i.e., child, parent, spouse, sibling, or other). Multivariate regression analyses were then used to predict the likelihood of smoking cessation and relapse controlling for the presence of other household members and their smoking status, sociodemographic characteristics, number of cigarettes smoked per day, previous quit attempts, and years abstained from smoking.

**Findings:** Individuals that lived with non-smokers were more likely to quit [OR1.22 (95%CI 1.11;1.34)] relative to those living alone. However, this favourable association was negated if living with another smoker, which was associated with a reduced likelihood of smoking cessation [OR0.77 (95%CI 0.72;0.83)] and a higher likelihood of relapse [1.37 (95%CI 1.22;1.53)]. In particular, living with a spouse or parent that smoked reduced the likelihood of smoking cessation [OR0.71 (95%CI 0.65;0.78) and OR0.71 (95%CI 0.59;0.84), respectively] and increased the likelihood of relapse [OR1.47 (95%CI 1.28;1.69) and OR1.39 (95%CI 1.00;1.94) respectively] relative to living with their non-smoking counterparts.

**Conclusions:** Household composition and intrahousehold smoking behaviour should be considered when delivering, or estimating the benefits of, smoking cessation interventions. Interventions which encourage smoking cessation at the household level may assist individuals to quit and abstain from smoking.

## Introduction

Tobacco smoking remains a public health challenge despite a significant decline in the prevalence of smoking in many countries over the past few decades (1). Whilst there is good evidence for the effectiveness of interventions to prompt and support people to quit smoking (2, 3), long-term quit rates remain low.

Although tobacco smoking is widely accepted as a social activity (4, 5), the influence of household members and their smoking behaviour on individuals’ smoking patterns has only briefly been explored in the literature. In the early 1980s and 90s, the absence of other household smokers was shown to be a strong predictor of smoking cessation among community samples (6-9). Christakis and Fowler (10) showed, using a sample from the US Framingham Heart Study that smoking cessation by a spouse or sibling reduced an individual’s likelihood of smoking. In the UK, women identified as smokers through the National Breast Screening Programme (aged 50-64 years), were more likely to quit smoking if they were partnered but less likely if that partner smoked (11). Another analysis of twins in the Netherlands found that smoking parents, siblings, or friends, increased individuals’ relative risk of smoking (12). A cross-sectional sample of adolescents in the US who lived with smoking parents were more likely to smoke compared to those with parents who had already given up smoking (13). Finally, in a representative and longitudinal British sample, members within the same household were observed to exhibit similar quitting smoking behaviours (14).

Generally, these studies have relied on cross-sectional samples or longitudinal cohorts with short-term, or infrequent, follow-up, which cannot account for the dynamics of smoking behaviour. Relapse has also been infrequently investigated. These dynamics are important as individuals often make multiple quit attempts during their lifetime and transition between periods of abstinence and relapse (15). Further, time-varying factors can influence an individual’s smoking behaviour, such as the socio-political context (e.g., taxation on tobacco products, media campaigns), duration of smoking abstinence, or indeed, peers’ smoking behaviour. Models, which do not capture the patterns of smoking cessation attempts over the long-term, can therefore bias the expected effectiveness of different interventions (16). Moreover, with many of these studies being published prior to 2010 or drawing evidence from non-representative populations, these models are unlikely to be suitable to inform current interventions.

Addressing this gap in the contemporary literature, this study investigates the potential mediating role of household composition and intrahousehold smoking behaviour on smoking cessation and relapse patterns among a nationally representative longitudinal cohort of Australians.

## Methods

The data used in this study is sourced from waves 1 to 19 of the Household Income and Labour Dynamics in Australia (HILDA) survey. HILDA is a national longitudinal household survey of over 17,000 households and has been conducted annually since 2001. HILDA has been described in more detail previously (17). HILDA collects information on various social, economic and wellbeing measures, with all household members aged 15 or older completing the survey. The data also contains detailed information about household structure and relationship types, which enables identification of respondents who live with their parents, spouse, children, siblings, or others in each wave.

The sample of smokers and ex-smokers were constructed based on the responses to the question: “Do you smoke cigarettes or any other tobacco products?” This question was asked in each wave. In wave 1, individuals were provided with three options: “No, I have never smoked”, “No, I have given up smoking,” and “Yes.” Respondents who selected the two last responses were considered to be ‘ex-smokers’ and ‘smokers’ respectively. In subsequent waves, respondents were again asked about their smoking status, albeit with the following response options: “No, I have never smoked”, “No, I no longer smoke”, “Yes, I smoke daily”, “Yes, I smoke at least weekly (but not daily)” and “Yes, I smoke less often than weekly.” Respondents were considered to be ‘smokers’ if they selected any of the last three choices (The proportion of smokers who selected each of these choices is reported in the supplementary materials SM.1). Applying this classification, the study sample was restricted to individuals who ever reported to be smoking throughout waves 1 to 19 (n=9,956) as well as those who reported they had “given up smoking” in the first wave (n=3,421). Although the question in wave 1 is useful for us to identify and include ‘ex-smokers’ in our sample, as the question and response options were different between wave 1 and subsequent waves, analyses were restricted to responses from wave 2 to wave 19. This produced a final sample of 12,723 unique individuals (noting there was a sample top-up in wave 11 (18)) who were classified, at some stage, as being a smoker or ex-smoker, with an average of 3,056 ex-smokers and 2,612 smokers per wave (Supplementary Material SM.2 provides the number of ex-smokers and smokers, and those with missing smoking status, for each wave).

Individuals were considered to have experienced a ‘quit event’ if they reported to be smoking in the previous year of the survey but reported were not smoking in the current year. If they were smoking in the previous year as well as the current year, they were considered to be still smoking. Following previous approaches (14), respondents were not included if their smoking status was missing in the current or previous year. Relapse events were defined similarly; if the individual was observed to have a quit event but then reported smoking in a subsequent wave, they were considered to have relapsed. Respondents with missing smoking status in the current year, or intervening years since their most recent quit attempt, were excluded from the relapse analyses.

Following previous studies in this space (19-21), a logistic model to predict the probability of quitting (for current smokers) or relapsing (for current ex-smokers) conditional on covariates was adopted. The quit and relapse models are shown in Model 1 and Model 2 respectively.

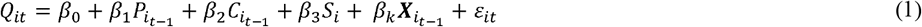

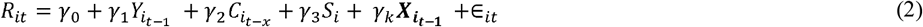

In Model 1, *Q*_*it*_ is a binary variable equal indicating whether individual *i* who reported to be a smoker in wave *t*-1 and a non-smoker in wave *t. P*_*it-*1_ is a binary indicator for whether they had previously reported being an ex-smoker. In Model 2, *R*_*it*_ is a binary variable indicating whether individual *i* who was an ex-smoker in wave *t-* 1 had relapsed in wave *t*, and 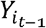 is a categorical variable indicating the time abstained from smoking (namely, 1-2, 2-3, 3-4, 4-7, or 7 or more consecutive waves without smoking). Following previous literature (22, 23), *C*_*i*_ is a categorical variable which classifies the individual as a ‘low’ (10 or fewer cigarettes per day), moderate (more than 10 and less than 20 cigarettes per day), or heavy smoker (20 or more cigarettes per day) and is based on the number of cigarettes an individual usually smoked per day in the year when they were last observed smoking (*t-* 1 for a quit event in Model 1 or *t-x* for a relapse event where *x* represents the number of years when the individual was last observed smoking). Both models control for sex of respondent (*S*_*i*_) and a vector of time varying individual level controls which are based on an individual’s responses in the previous wave (*X*_*it-*1_); namely, age group; level of education (below high school, high school, or university degree); and quartile of equivalised household income.

Subsequent to these main specifications, the role of within-household social support and smoking behaviour was investigated. First, covariates indicating whether the individual was living with another person in the previous wave, and whether that person was smoking, were included (household members under 15 years old are used to determine household structure, however, their smoking status is unobserved). Next, the presence of household members, and their smoking status, was disaggregated by their relationship to the individual. Specifically, for the latter, whether smoking patterns vary depending on whether the other household member is a parent, spouse, sibling, child, or other household member was explored. This flexible structure allowed for the association to change for different household members and for the combined effects of different household structures to be estimated. To facilitate the interpretation of the model outputs, the average predicted smoking cessation and relapse rates were estimated for individuals living with different smoking and non-smoking household members, holding all other covariates at their observed values.

Multiple robustness checks were conducted. First, different specifications were applied which classified individuals who smoked weekly (but not daily), as well as those that smoked less often than weekly, as non-smokers. Next, although HILDA has relatively low attrition rates and was supplemented with a sample top-up in wave 11 (18, 24), the stability of our estimates over time was explored by re-estimating the main specification models with year fixed effects (including dummy variables for each year) and through limiting the sample to the last five years (i.e., between 2014 to 2019). Third, as HILDA is asked at one time point in any given year, individuals might have only recently quit (for instance, within two weeks prior to the survey). The association of intrahousehold peer effects with ‘sustained’ quit attempts was then explored by re-classifying a quit attempt as two consecutive waves without smoking. Finally, to test whether concern for smoke exposure to children is a potential motivator to quit and abstain from smoking, the associations between household members and smoking behaviour was disaggregated by whether household members were under the age of 15 years.

All regression analyses were performed in Stata version 16 (25) with standard errors robust clustered at the individual level.

## Results

### Descriptive statistics

The descriptive characteristics at the mid-point of the observed sample in 2008 (wave 8) sample of smokers and ex-smokers are shown in Table 1 below. In 2008, there were 2,725 smokers and 2,247 ex-smokers in the sample. Compared to those who were currently smoking, ex-smokers were older (mean 53 vs 40 years, p<0.001), had higher household incomes and more years of education. When they were last observed smoking, ex-smokers were more likely to have been smoking fewer cigarettes. Of the smokers, 15% reported they were not smoking in the subsequent wave (i.e., quit the following year) while 6% of ex-smokers reported they were smoking again in the subsequent wave (i.e., relapsed). Current smokers had slightly more people living with them compared to ex-smokers (mean 2.67 vs 2.83, p<0.001) and were more likely to live with siblings (mean 0.09 vs 0.02, p<0.001), parents (mean 0.13 vs 0.04, p<0.001), and other household members (mean 0.10 vs 0.05, p<0.001). A smaller proportion of current smokers lived with a partner compared to ex-smokers (mean 0.56 vs 0.72, p<0.001) while the proportion of individuals living with children was similar across both groups (mean 0.40 vs 0.40, p=0.96).

**Table 1.** Descriptive characteristics of wave 8 respondents (year 2008)

|  | Not currently smoking<br>(n=2,725) |  | Currently smoking<br>(n=2,247) |  |
| --- | --- | --- | --- | --- |
|  | Mean/Prop | Freq. | Mean/Prop | Freq. |
| ***Outcomes*** |  |  |  |  |
| Quit in next year | - | - | 0.15 | 275 |
| Relapse in next year | 0.06 | 146 | - | - |
| ***Characteristics*** |  |  |  |  |
| Male | 0.50 | 1,376 | 0.53 | 1,182 |
| ***Age group*** |  |  |  |  |
| Under 30 | 0.09 | 239 | 0.31 | 687 |
| 30-39 | 0.13 | 357 | 0.20 | 455 |
| 40-49 | 0.22 | 608 | 0.23 | 509 |
| 50-59 (Ref) | 0.20 | 535 | 0.16 | 350 |
| 60-69 | 0.19 | 518 | 0.08 | 175 |
| Over 70 | 0.17 | 468 | 0.03 | 71 |
| ***Educational attainment*** |  |  |  |  |
| Less than high school | 0.33 | 901 | 0.41 | 917 |
| High school or equivalent | 0.34 | 926 | 0.42 | 935 |
| University degree or above | 0.33 | 896 | 0.18 | 394 |
| ***Quartile of equivalised household income*** |  |  |  |  |
| First quartile (lowest income) | 0.28 | 754 | 0.28 | 626 |
| Second quartile | 0.22 | 586 | 0.26 | 578 |
| Third quartile | 0.24 | 655 | 0.26 | 580 |
| Fourth quartile (highest income) | 0.27 | 730 | 0.21 | 463 |
| Previous quit recorded | 0.94 | 2,556 | 0.25 | 565 |
| ***Level of smoking:*** |  |  |  |  |
| 10 or fewer cigarettes per day | 0.73 | 633 | 0.55 | 1,224 |
| 11-19 cigarettes per day | 0.14 | 124 | 0.24 | 529 |
| 20 or more cigarettes per day | 0.13 | 116 | 0.21 | 472 |
| Number of people in household | 2.67 | 7,271 | 2.83 | 6,354 |
| Living with partner | 0.72 | 1,956 | 0.56 | 1,248 |
| Living with child | 0.40 | 1,099 | 0.40 | 905 |
| Living with child – under 15 | 0.29 | 786 | 0.31 | 694 |
| Living with parent | 0.04 | 115 | 0.13 | 298 |
| Living with sibling | 0.02 | 66 | 0.09 | 203 |
| Living with other household member | 0.05 | 124 | 0.10 | 225 |
| Smoker in household | 0.13 | 354 | 0.39 | 884 |
| Number of smokers in household | 0.14 | 393 | 0.45 | 1,008 |
| Living with smoking partner | 0.08 | 217 | 0.27 | 596 |
| Living with smoking child | 0.04 | 120 | 0.06 | 125 |
| Living with smoking parent | 0.00 | 11 | 0.05 | 106 |
| Living with smoking sibling | 0.00 | 10 | 0.03 | 59 |
| Living with other smoking household member | 0.01 | 22 | 0.04 | 83 |

**Note:** All characteristics are based on wave 8 responses. Those who had not yet started smoking (i.e., as of wave 8, had never smoked) are not included in this table (n=555).

A significantly higher proportion of current smokers lived with another smoker in their household compared to ex-smokers (mean 0.39 vs 0.13, p<0.001) and the number of other smokers in their household was higher (mean 0.45 vs 0.14, p<0.001). Compared to ex-smokers, the proportion of household members that smoked was higher across all relationship types; current smokers were more likely to live with a partner, child, parent, sibling, or other household member that smoked.

Observed characteristics are similar to the 2018 sample (Supplementary Material SM.3).

### Smoking cessation analysis

The regression analyses results predicting the likelihood of quitting smoking are shown in Table 2. The first column presents the baseline model specification without any household member information, the second column includes the baseline covariates as well as information on whether they live with another household member and their smoking status, the third column includes the baseline covariates as well as information on relationship with cohabiting household members and their smoking status.

**Table 2.** **Smoking cessation models adjusting for (1) baseline covariates; (2) baseline covariates, cohabitant smoking status; (3) baseline covariates, cohabitant smoking status by relationship**.

|  | (1)<br>Quit<br>OR | [95% CI] | (2)<br>Quit<br>OR | [95% CI] | (3)<br>Quit<br>OR | [95% CI] |
| --- | --- | --- | --- | --- | --- | --- |
| Previous quit recorded | 2.24*** | [2.08;2.41] | 2.22*** | [2.06;2.39] | 2.24*** | [2.08;2.42] |
| *Level of smoking:* |  |  |  |  |  |  |
| 10 or fewer cigarettes per day<br>(Ref) | 1.00 | [1.00;1.00] | 1.00 | [1.00;1.00] | 1.00 | [1.00;1.00] |
| 11-19 cigarettes per day | 0.55*** | [0.50;0.59] | 0.55*** | [0.51;0.60] | 0.55*** | [0.51;0.60] |
| 20 or more cigarettes per day | 0.46*** | [0.42;0.51] | 0.47*** | [0.43;0.52] | 0.48*** | [0.43;0.53] |
| *Sex:* |  |  |  |  |  |  |
| Male | 0.99 | [0.93;1.07] | 0.99 | [0.92;1.06] | 0.95 | [0.89;1.02] |
| *Age (years):* |  |  |  |  |  |  |
| Under 30 | 1.59*** | [1.43;1.77] | 1.62*** | [1.45;1.80] | 1.66*** | [1.48;1.86] |
| 30-39 | 1.25*** | [1.12;1.40] | 1.25*** | [1.12;1.39] | 1.32*** | [1.18;1.48] |
| 40-49 | 0.98 | [0.88;1.10] | 0.98 | [0.88;1.09] | 1.02 | [0.92;1.14] |
| 50-59 (Ref) | 1.00 | [1.00;1.00] | 1.00 | [1.00;1.00] | 1.00 | [1.00;1.00] |
| 60-69 | 1.33*** | [1.16;1.53] | 1.31*** | [1.14;1.51] | 1.27*** | [1.10;1.45] |
| Over 70 | 1.32** | [1.07;1.63] | 1.29* | [1.04;1.59] | 1.22 | [0.99;1.51] |
| *Education:* |  |  |  |  |  |  |
| Less than high school | 1.00 | [1.00;1.00] | 1.00 | [1.00;1.00] | 1.00 | [1.00;1.00] |
| High school or equivalent | 1.15*** | [1.06;1.24] | 1.14** | [1.05;1.23] | 1.14** | [1.05;1.23] |
| University degree | 1.43*** | [1.30;1.57] | 1.42*** | [1.29;1.56] | 1.41*** | [1.28;1.56] |
| *Equivalised household income:* |  |  |  |  |  |  |
| First quartile (lowest income) | 1.00 | [1.00;1.00] | 1.00 | [1.00;1.00] | 1.00 | [1.00;1.00] |
| Second quartile | 1.07 | [0.98;1.17] | 1.06 | [0.97;1.16] | 1.05 | [0.96;1.15] |
| Third quartile | 1.24*** | [1.13;1.36] | 1.23*** | [1.12;1.35] | 1.18*** | [1.08;1.30] |
| Fourth quartile (highest income) | 1.50*** | [1.36;1.65] | 1.47*** | [1.34;1.62] | 1.39*** | [1.26;1.53] |
| *Household:* |  |  |  |  |  |  |
| Lives with non-smoker | - | - | 1.22*** | [1.11;1.34] | - | - |
| Lives with smoker | - | - | 0.77*** | [0.72;0.83] | - | - |
| Lives with non-smoking spouse | - | - | - | - | 1.34*** | [1.23;1.46] |
| Lives with non-smoking child | - | - | - | - | 0.88** | [0.81;0.96] |
| Lives with non-smoking parent | - | - | - | - | 1.02 | [0.89;1.18] |
| Lives with non-smoking sibling | - | - | - | - | 1.26** | [1.07;1.48] |
| Lives with other non-smoker | - | - | - | - | 1.12 | [0.98;1.27] |
| Lives with smoking spouse | - | - | - | - | 0.71*** | [0.65;0.78] |
| Lives with smoking child | - | - | - | - | 1.02 | [0.87;1.19] |
| Lives with smoking parent | - | - | - | - | 0.71*** | [0.59;0.84] |
| Lives with smoking sibling | - | - | - | - | 0.83 | [0.67;1.03] |
| Lives with other smoker | - | - | - | - | 0.80* | [0.66;0.98] |
| Constant | 0.10*** | [0.09;0.11] | 0.10*** | [0.08;0.11] | 0.10*** | [0.09;0.11] |
| N | 37,096 |  | 37,096 |  | 37,096 |  |
| Mean of outcome | 0.14 |  | 0.14 |  | 0.14 |  |

**Note:** \*\*\*p < 0.01; \*\*p < 0.05; \*p < 0.10. With the exception of past level of smoking, which was recorded when the individual was last observed to be smoking, all characteristics are based on the individual/household information from the previous wave.

Compared to individuals living on their own, those living with at least one other non-smoker were more likely to quit [OR 1.22 (95%CI 1.11;1.34)]. However, this positive association did not hold if a cohabitant was also a smoker. Living with at least one other smoker lowered the odds of quitting OR 0.77 (95%CI 0.72;0.83). The association between the smoking household member and likelihood of smoking cessation also varied significantly depending on the relationship to the individual. Individuals were significantly less likely to quit if the other household smoker was their parent [OR 0.71 (95%CI 0.59;0.84)], spouse [OR 0.71 (95%CI 0.65;0.78)], or other household member [OR 0.80 (95%CI 0.66;0.98)].

Figure 1a shows the average predicted probability of smoking cessation for a smoker living alone and with different smoking or non-smoking household members for the sample using column 3 estimates. Across the whole sample, the true annual quit rate is 14.4%. The predicted annual proportion reporting smoking cessation for smokers living alone is 14.0% compared to 17.7%, 16.9% and 14.3%, for those living with a non-smoking partner, sibling, and parent respectively. Conversely, the quit rate is slightly lower among those living with a non-smoking child over 15 years old (12.6%). Compared to those living alone, the quit rate is lower for individuals living with a smoking parent (10.7%), a smoking child (12.9%), and a smoking partner (13.5%).

**Figure 1.**
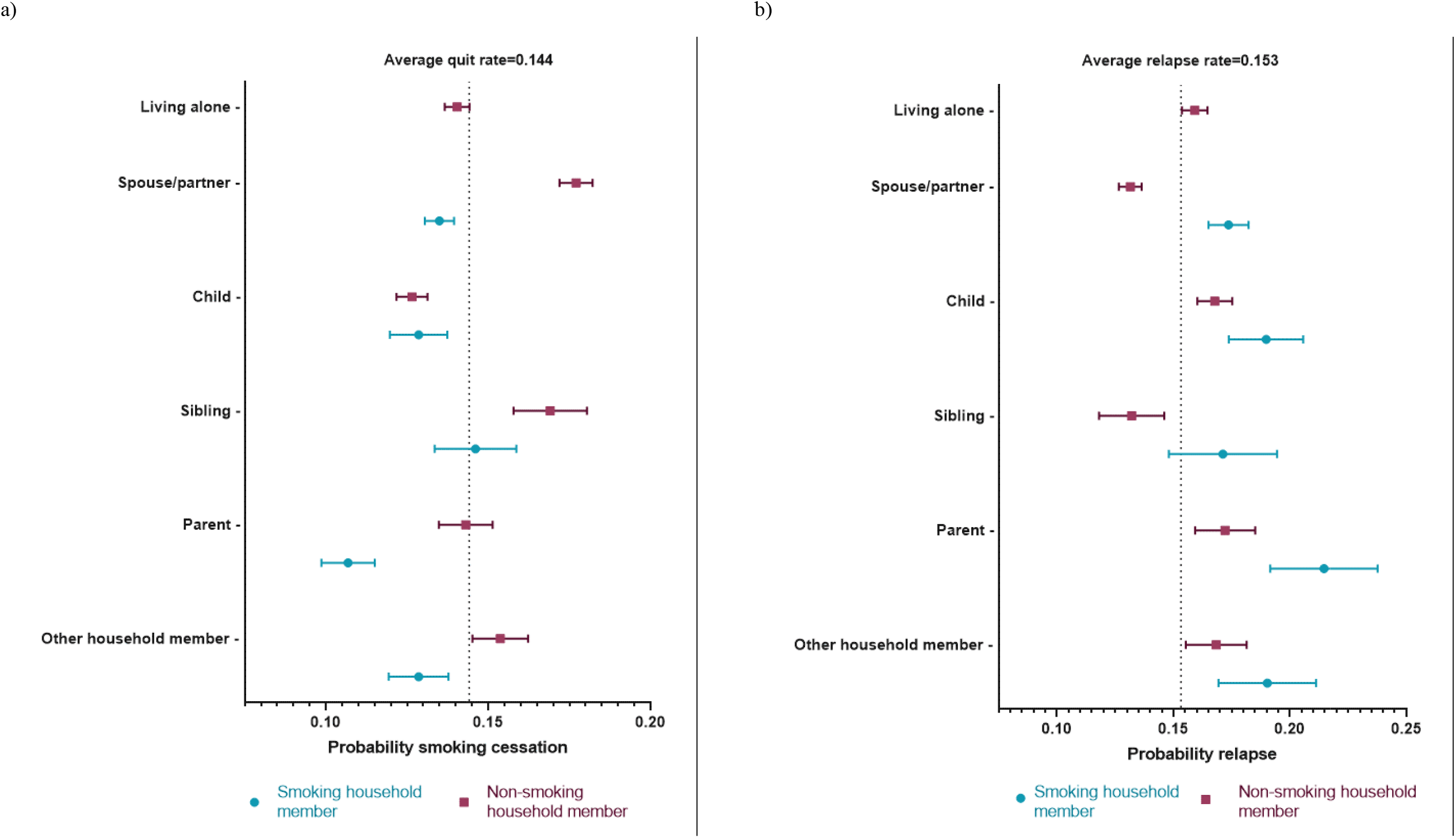
**Average predicted probability of smoking cessation (a) and relapse (b) for different types of household structure**. **Note: Applying estimates from smoking cessation and relapse models column 3 for the whole population. Average quit and relapse rates for sample population a
n at observed valu**

### Relapse analysis

The regression analyses results predicting the likelihood of relapse is shown in Table 3. As for the smoking cessation analysis, column 1 presents the baseline specification, the second adds whether they lived with another household member and if that household member smoked, and the third column includes information on the presence, and smoking status, of household members by their relationship to the individual.

**Table 3.** **Smoking relapse models adjusting for (1) baseline covariates; (2) baseline covariates, cohabitant smoking status; (3) baseline covariates, cohabitant smoking status by relationship**.

|  | (1)<br>Relapse<br>OR | [95% CI] | (2)<br>Relapse<br>OR | [95% CI] | (3)<br>Relapse<br>OR | [95% CI] |
| --- | --- | --- | --- | --- | --- | --- |
| *Consecutive waves abstained:* |  |  |  |  |  |  |
| 1-2 (ref) | 1.00 | [1.00;1.00] | 1.00 | [1.00;1.00] | 1.00 | [1.00;1.00] |
| 2-3 | 0.36*** | [0.32;0.40] | 0.36*** | [0.32;0.41] | 0.36*** | [0.32;0.41] |
| 3-4 | 0.21*** | [0.18;0.24] | 0.21*** | [0.18;0.25] | 0.21*** | [0.18;0.25] |
| 4-7 | 0.10*** | [0.09;0.12] | 0.10*** | [0.09;0.12] | 0.10*** | [0.09;0.12] |
| 7 or more | 0.04*** | [0.03;0.05] | 0.04*** | [0.03;0.05] | 0.04*** | [0.03;0.05] |
| *Level of smoking:* |  |  |  |  |  |  |
| 10 or fewer cigarettes per day<br>(Ref) | 1.00 | [1.00;1.00] | 1.00 | [1.00;1.00] | 1.00 | [1.00;1.00] |
| 11-19 cigarettes per day | 1.14* | [1.01;1.29] | 1.14* | [1.00;1.29] | 1.14* | [1.00;1.29] |
| 20 or more cigarettes per day | 1.06 | [0.92;1.23] | 1.05 | [0.91;1.22] | 1.05 | [0.91;1.22] |
| *Sex:* |  |  |  |  |  |  |
| Male | 1.04 | [0.95;1.14] | 1.06 | [0.97;1.16] | 1.09 | [0.99;1.20] |
| *Age (years):* |  |  |  |  |  |  |
| Under 30 | 1.19* | [1.03;1.38] | 1.18* | [1.02;1.37] | 1.14 | [0.97;1.33] |
| 30-39 | 1.06 | [0.91;1.23] | 1.06 | [0.91;1.23] | 1.06 | [0.91;1.24] |
| 40-49 | 1.09 | [0.93;1.26] | 1.08 | [0.93;1.25] | 1.07 | [0.91;1.25] |
| 50-59 (Ref) | 1.00 | [1.00;1.00] | 1.00 | [1.00;1.00] | 1.00 | [1.00;1.00] |
| 60-69 | 0.70*** | [0.58;0.84] | 0.71*** | [0.59;0.85] | 0.72*** | [0.60;0.87] |
| Over 70 | 0.52*** | [0.39;0.69] | 0.53*** | [0.40;0.71] | 0.55*** | [0.41;0.73] |
| *Education:* |  |  |  |  |  |  |
| Less than high school | 1.00 | [1.00;1.00] | 1.00 | [1.00;1.00] | 1.00 | [1.00;1.00] |
| High school or equivalent | 0.93 | [0.83;1.05] | 0.95 | [0.84;1.06] | 0.95 | [0.84;1.06] |
| University degree | 0.80** | [0.70;0.92] | 0.83** | [0.72;0.95] | 0.82** | [0.72;0.94] |
| *Equivalised household income:* |  |  |  |  |  |  |
| First quartile (lowest income) | 1.00 | [1.00;1.00] | 1.00 | [1.00;1.00] | 1.00 | [1.00;1.00] |
| Second quartile | 0.85* | [0.74;0.97] | 0.84* | [0.73;0.96] | 0.86* | [0.75;0.99] |
| Third quartile | 0.75*** | [0.66;0.86] | 0.74*** | [0.65;0.85] | 0.78*** | [0.68;0.89] |
| Fourth quartile (highest income) | 0.73*** | [0.63;0.83] | 0.72*** | [0.63;0.83] | 0.78*** | [0.67;0.90] |
| *Household:* |  |  |  |  |  |  |
| Lives with non-smoker | - | - | 1.01 | [0.89;1.15] | - | - |
| Lives with smoker | - | - | 1.37*** | [1.22;1.53] | - | - |
| Lives with non-smoking spouse | - | - | - | - | 0.77*** | [0.68;0.87] |
| Lives with non-smoking child | - | - | - | - | 1.08 | [0.96;1.21] |
| Lives with non-smoking parent | - | - | - | - | 1.12 | [0.90;1.40] |
| Lives with non-smoking sibling | - | - | - | - | 0.77 | [0.59;1.01] |
| Lives with other non-smoker | - | - | - | - | 1.08 | [0.87;1.34] |
| Lives with smoking spouse | - | - | - | - | 1.47*** | [1.28;1.69] |
| Lives with smoking child | - | - | - | - | 1.20 | [0.94;1.53] |
| Lives with smoking parent | - | - | - | - | 1.39* | [1.00;1.94] |
| Lives with smoking sibling | - | - | - | - | 1.43 | [0.97;2.11] |
| Lives with other smoker | - | - | - | - | 1.20 | [0.82;1.74] |
| Constant | 0.70*** | [0.60;0.83] | 0.64*** | [0.53;0.78] | 0.70*** | [0.59;0.84] |
| *N* | 17,478 |  | 17,478 |  | 17,478 |  |
| *Mean of outcome* | 0.15 |  | 0.15 |  | 0.15 |  |

**Note:** \*\*\*p < 0.01, \*\*p < 0.05, \*p < 0.10. With the exception of past level of smoking, which was recorded when the individual was last observed to be smoking, all characteristics are based on the individual/household information from the previous wave.

Compared to those living alone, individuals living in households with non-smokers were not any more likely to relapse [OR 1.01 (95%CI 0.89;0.1.15)] but the presence of at least one other smoker in the household was associated with significantly higher odds of relapse [OR 1.37 (95%CI 1.22;1.53)]. Across all relationship types, cohabiting with a spouse who smoked was the strongest predictor of relapse [OR 1.47 (1.28;1.69)], followed by living with a smoking parent [OR 1.39 95%CI (1.00;1.94)]. Living with a smoking child, sibling, or other household smoker were all positively associated with relapse but were statistically insignificant.

The predicted probability of relapse for an individual living alone, and with different smoking or non-smoking household members, based on column 3 estimates, is presented in Figure 1b. Across the whole sample, the true annual relapse rate is 15.3%. The annual proportion reporting smoking relapse for ex-smokers living alone is 15.9% compared to 13.1% and 13.2%, for those living with a non-smoking partner and non-smoking sibling, respectively. Conversely, the relapse rate is slightly higher among those living with a non-smoking child, parent or other non-smoking household member (16.8, 17.2 and 16.8% respectively). The relapse rate individuals living with a smoking parent, a smoking child, and a smoking partner is 21.5%, 19.0%, and 17.4% respectively.

### Robustness checks

The smoking cessation and relapse models were robust to multiple specifications and subgroup analyses. The estimates were relatively stable when restricting the sample to the most recent 5 waves and adding wave fixed effects (Supplementary Material SM.4). The presence of a smoking parent or partner was still positively associated with relapse, with similar point estimates, when restricting the sample to the most recent 5 waves as well as including wave fixed effects. Including wave fixed effects did not change the level of significance nor direction of the associations for any of the intrahousehold smoking behaviour covariates. When including only waves between 2015-2019 however, the presence of a smoking spouse, parent, or other household member was still positively associated with smoking cessation, but the estimates were no longer statistically significant. Classifying those who reported that they smoked less than weekly as well as those that described themselves as smoking weekly but less than daily (Supplementary Material SM.5) as non-smokers did not substantially change the associations nor our conclusions and only dampened the association between the presence of a smoking parent and reduced odds of smoking cessation. Reclassifying quit attempts as two consecutive waves without smoking (Supplementary Material SM.6), strengthened the negative association with the presence of smoking partners, parents, and other household members on smoking cessation. Lastly, living with household members that were under 15 was not significantly associated with smoking cessation nor relapse (Supplementary Material SM.7). The association between living with a child and being less likely to quit was mainly attributable to those individuals living with children aged 15 and above.

## Discussion

This study investigated the association between household structure and intrahousehold smoking behaviour on smoking cessation and relapse patterns among a longitudinal and representative cohort. Our results indicate that living with non-smokers is generally associated with increased likelihood of smoking cessation and reduced likelihood of relapse while the presence of other smokers in the household has the opposite associations. In particular, living with a smoking parent or spouse was associated with reduced odds of smoking cessation and increased odds of relapse.

Our findings are generally consistent with previous work exploring the influence of other family members (5, 9, 11, 14, 26, 27) and their smoking behaviour (10, 13, 28) on individuals’ smoking cessation and relapse. We build upon this research by providing a population-based model which accounts for the dynamic nature of explanatory variables with annual follow-up over a substantially longer observation period of 19 years. Moreover, this study provides comprehensive evidence about how smoking patterns vary by different relationships within the household. Importantly, these findings suggest that intrahousehold connections can have both positive and negative influences on smoking behaviour. We show that compared to individuals living alone, living with others, particularly non-smoking partners and siblings, is associated with increased odds of smoking cessation. This aligns with research that other household members can encourage or support smoking cessation and abstinence (27) as well as broader research suggesting that social support is important for improving health behaviours (29). Conversely, our findings that living with other smokers, in particular smoking parents or spouses, is associated with reduced odds of smoking cessation and abstinence, reinforces that decisions to quit, and abstain from, cigarette smoking are not made in isolation.

These results have direct implications for policy and future modelling. While the current evidence base suggests that social support interventions, such as interventions to enhance partner support, do not increase long-term quit rates (27), our findings highlight household structure and smokers in the household are nevertheless important predictors of an individual’s smoking status. This suggest that interventions which may encourage simultaneous smoking cessation or relapse avoidance at the household, or broader societal, level, such as smoke-free legislation or cigarette taxes, may be more effective at affecting smoking behaviour than interventions targeted to specific individuals or households. More specifically, evaluations which neglect the impact of peers on smoking behaviour in the long-term may over-estimate the benefits of smoking cessation interventions. The estimates presented herein provide a useful tool for future modelling and cost effectiveness analyses in smoking cessation trials.

The results of this study should be interpreted within the context of several limitations. First, given the HILDA survey is conducted once per year, we do not have definitive measures of quit, relapse, nor abstinence rates at more frequent intervals. However, given we show the presence of household smokers was still significantly associated with reduced odds of a sustained quit events (abstaining for 1-2 years), this provides us with some confidence that intrahousehold peer effects are important for both shorter-term and longer-term quit attempts. Second, we do not observe whether quit attempts were facilitated by smoking cessation aids or other formal behavioural support, which has shown to lead to more sustained quit attempts relative to ‘cold turkey’ cessation (30-32). The self-reported nature of smoking status may also be subject to measurement error. However, previous research has demonstrated that self-report measures are generally consistent with biochemically validated measures (33, 34). Finally, data limitations pertaining to sample size and omission of questions pertaining to the smoking status of other peers, such as close friends, or co-workers, imposes limitations and hinders our ability to understand which peers are important across different stages of the life course. Capturing robust measures of intrahousehold and broader peers’ smoking behaviours in large national samples is recommended to facilitate understanding around which groups should be targeted at different ages.

Despite these limitations, these results provide dynamic models of smoking cessation and relapse and ultimately highlight the importance of peer effects on individual smoking patterns. The long-term follow up of smokers and ex-smokers from a nationally representative and contemporary sample further bolsters the model’s external validity and suitability to inform public health interventions. These findings may also have implications for other alcohol and drug treatment programs where other family factors would merit consideration.

## Data Availability

All data produced in the present study are available upon reasonable request to the Australian Government Department of Social Services

## Acknowledgements

This research was supported by project grants 1108318, 1127390, 1162583, and career development fellowship grant 1148497 (awarded to Dr Courtney) from the Australian National Health and Medical Research Council.

