## Supplementary Material for "Household composition, smoking cessation and relapse: results from a prospective longitudinal Australian cohort"

### Supplementary Materials

#### SM.1. Daily, weekly and less than weekly smokers by wave

| **Wave** | **Smokes daily** | **Smokes at least weekly  (but not daily)** | **Smokes less often than weekly** | **Total** |
| --- | --- | --- | --- | --- |
| 2 | 2,368 | 296 | 237 | 2,901 |
|  | *81.63* | *10.2* | *8.17* | *100* |
| 3 | 2,266 | 264 | 204 | 2,734 |
|  | *82.88* | *9.66* | *7.46* | *100* |
| 4 | 2,136 | 267 | 194 | 2,597 |
|  | *82.25* | *10.28* | *7.47* | *100* |
| 5 | 2,101 | 264 | 211 | 2,576 |
|  | *81.56* | *10.25* | *8.19* | *100* |
| 6 | 2,016 | 255 | 186 | 2,457 |
|  | *82.05* | *10.38* | *7.57* | *100* |
| 7 | 1,958 | 229 | 184 | 2,371 |
|  | *82.58* | *9.66* | *7.76* | *100* |
| 8 | 1,879 | 179 | 189 | 2,247 |
|  | *83.62* | *7.97* | *8.41* | *100* |
| 9 | 1,853 | 220 | 189 | 2,262 |
|  | *81.92* | *9.73* | *8.36* | *100* |
| 10 | 1,965 | 208 | 194 | 2,367 |
|  | *83.02* | *8.79* | *8.2* | *100* |
| 11 | 2,369 | 255 | 266 | 2,890 |
|  | *81.97* | *8.82* | *9.2* | *100* |
| 12 | 2,244 | 277 | 270 | 2,791 |
|  | *80.4* | *9.92* | *9.67* | *100* |
| 13 | 2,184 | 286 | 238 | 2,708 |
|  | *80.65* | *10.56* | *8.79* | *100* |
| 14 | 2,257 | 277 | 238 | 2,772 |
|  | *81.42* | *9.99* | *8.59* | *100* |
| 15 | 2,145 | 283 | 219 | 2,647 |
|  | *81.04* | *10.69* | *8.27* | *100* |
| 16 | 2,304 | 312 | 240 | 2,856 |
|  | *80.67* | *10.92* | *8.4* | *100* |
| 17 | 2,129 | 323 | 239 | 2,691 |
|  | *79.12* | *12* | *8.88* | *100* |
| 18 | 2,033 | 274 | 257 | 2,564 |
|  | *79.29* | *10.69* | *10.02* | *100* |
| 19 | 2,037 | 305 | 237 | 2,579 |
|  | *78.98* | *11.83* | *9.19* | *100* |

#### SM.2. Number of smokers and ex-smokers for each wave

The number of smokers and ex-smokers observed across each of the waves is presented below. These figures exclude those who have not been observed to be smoking yet.

| **Wave** | **Ex-smoker** | **Smoker** | **Smoking status  missing** | **Total** |
| --- | --- | --- | --- | --- |
| 2 | 2,889 | 2,901 | 418 | 6,208 |
| 3 | 2,829 | 2,734 | 503 | 6,066 |
| 4 | 2,742 | 2,597 | 519 | 5,858 |
| 5 | 2,737 | 2,576 | 672 | 5,985 |
| 6 | 2,833 | 2,457 | 662 | 5,952 |
| 7 | 2,794 | 2,371 | 726 | 5,891 |
| 8 | 2,725 | 2,247 | 856 | 5,828 |
| 9 | 2,823 | 2,262 | 967 | 6,052 |
| 10 | 2,958 | 2,367 | 732 | 6,057 |
| 11 | 2,986 | 2,890 | 956 | 6,832 |
| 12 | 3,082 | 2,791 | 994 | 6,867 |
| 13 | 3,213 | 2,708 | 973 | 6,894 |
| 14 | 3,259 | 2,772 | 855 | 6,886 |
| 15 | 3,298 | 2,647 | 945 | 6,890 |
| 16 | 3,406 | 2,856 | 634 | 6,896 |
| 17 | 3,477 | 2,691 | 668 | 6,836 |
| 18 | 3,450 | 2,564 | 696 | 6,710 |
| 19 | 3,512 | 2,579 | 592 | 6,683 |
| *Total* | *55,013* | *47,010* | *13,368* | *115,391* |

The number of individuals who are yet to be observed smoking (and did not describe themselves as an ex-smoker in wave 1) are presented below for each wave.

| **Wave** | **Observed smoking** | **Not yet observed smoking** |
| --- | --- | --- |
| 2 | 2,674 | 208 |
| 3 | 2,559 | 154 |
| 4 | 2,439 | 120 |
| 5 | 2,449 | 89 |
| 6 | 2,376 | 71 |
| 7 | 2,278 | 51 |
| 8 | 2,234 | 36 |
| 9 | 2,242 | 28 |
| 10 | 2,177 | 25 |
| 11 | 2,128 | 20 |
| 12 | 2,066 | 15 |
| 13 | 2,035 | 11 |
| 14 | 2,000 | 12 |
| 15 | 1,970 | <10 |
| 16 | 1,936 | <10 |
| 17 | 1,906 | <10 |
| 18 | 1,852 | <10 |
| 19 | 1,800 | <10 |
| *Total* | *39,121* | *859* |

#### SM.3. Descriptive characteristics of wave 18 respondents (year 2018)

|  | Not currently smoking (n=3,450) | | Currently smoking (2,564) | |
| --- | --- | --- | --- | --- |
|  | Mean/Prop. | Freq. | Mean/Prop. | Freq. |
| Quit in next year |  |  | .16 | 346 |
| Relapse in next year | 0.07 | 213 |  |  |
| Male | 0.51 | 1749 | 0.53 | 1371 |
| *Age group* |  |  |  |  |
| Age: <30 | 0.09 | 325 | 0.28 | 706 |
| [30-40) | 0.16 | 554 | 0.21 | 533 |
| [40-50) | 0.14 | 491 | 0.19 | 494 |
| [50-60) | 0.20 | 707 | 0.19 | 476 |
| [60-70) | 0.19 | 655 | 0.10 | 245 |
| >=70 | 0.21 | 718 | 0.04 | 110 |
| *Educational attainment* |  |  |  |  |
| less than Yr 12 | 0.25 | 861 | 0.32 | 824 |
| Yr 12 or equivalent | 0.39 | 1339 | 0.48 | 1242 |
| Bachelor or above | 0.36 | 1248 | 0.19 | 497 |
| *Quartile of equivalised household income* |  |  |  |  |
| 1 | 0.28 | 979 | 0.33 | 837 |
| 2 | 0.21 | 736 | 0.29 | 737 |
| 3 | 0.25 | 860 | 0.22 | 563 |
| 4 | 0.25 | 875 | 0.17 | 427 |
| Previous quit recorded | 0.94 |  | 0.34 |  |
| *No. cigarettes smoked per day* |  |  |  |  |
| No. cigs smoked per day: <=10 | 0.73 | 1449 | 0.60 | 1525 |
| (10-25) | 0.16 | 312 | 0.23 | 583 |
| >=20 | 0.11 | 220 | 0.17 | 421 |
| Number of people in household | 2.58 | 8898 | 2.85 | 7320 |
| Living with partner | 0.69 | 2392 | 0.53 | 1348 |
| Living with child | 0.39 | 1332 | 0.42 | 1065 |
| Living with parent | 0.04 | 145 | 0.13 | 340 |
| Living with sibling | 0.02 | 86 | 0.09 | 233 |
| Living with other household member | 0.05 | 177 | 0.12 | 306 |
| Smoker in household | 0.14 | 477 | 0.47 | 1194 |
| Number of smokers in household | 0.12 | 422 | 0.39 | 990 |
| Living with smoking partner | 0.08 | 259 | 0.25 | 638 |
| Living with smoking child | 0.03 | 101 | 0.06 | 146 |
| Living with smoking parent | 0.01 | 34 | 0.06 | 142 |
| Living with smoking sibling | 0.01 | 28 | 0.03 | 74 |
| Living with other smoking household member | 0.01 | 34 | 0.05 | 122 |

**Note: Characteristics are based on wave 18, not wave 17, information.**

#### SM.4. Restricting main specification to 2014 to 2019 and including wave fixed effects

Table A. 1 – Smoking cessation models when restricting sample to 2014 to 2019 (1) and including year fixed effects (2)

|  | (1) |  | (2) |  |
| --- | --- | --- | --- | --- |
|  | **Quit** |  | **Quit** |  |
|  | $OR$ | [95% CI] | $OR$ |  |
| Previous quit recorded | 2.17^***^ | [1.93,2.44] | 2.25^***^ | [2.08,2.42] |
| *Level of smoking:* |  |  |  |  |
| 10 or fewer cigarettes per day (Ref) | 1.00 | [1.00,1.00] | 1.00 | [1.00,1.00] |
| 11-19 cigarettes per day | 0.54^***^ | [0.46,0.62] | 0.55^***^ | [0.51,0.60] |
| 20 or more cigarettes per day | 0.46^***^ | [0.38,0.55] | 0.47^***^ | [0.43,0.52] |
| *Sex:* |  |  |  |  |
| Male | 1.01 | [0.90,1.13] | 0.95 | [0.89,1.02] |
| *Age (years):* |  |  |  |  |
| Under 30 | 1.74^***^ | [1.44,2.09] | 1.67^***^ | [1.49,1.87] |
| 30-39 | 1.43^***^ | [1.19,1.72] | 1.33^***^ | [1.19,1.49] |
| 40-49 | 0.90 | [0.74,1.10] | 1.03 | [0.92,1.15] |
| 50-59 (Ref) | 1.00 | [1.00,1.00] | 1.00 | [1.00,1.00] |
| 60-59 | 1.31^*^ | [1.05,1.64] | 1.27^***^ | [1.10,1.46] |
| Over 70 | 0.96 | [0.67,1.38] | 1.22 | [0.99,1.52] |
| *Education:* |  |  |  |  |
| Less than high school | 1.00 | [1.00,1.00] | 1.00 | [1.00,1.00] |
| High school or equivalent | 1.11 | [0.97,1.27] | 1.14^**^ | [1.05,1.23] |
| University degree | 1.29^**^ | [1.09,1.52] | 1.41^***^ | [1.28,1.56] |
| *Equivalised household income:* |  |  |  |  |
| First quartile (lowest income) | 1.00 | [1.00,1.00] | 1.00 | [1.00,1.00] |
| Second quartile | 1.14 | [0.98,1.33] | 1.05 | [0.96,1.15] |
| Third quartile | 1.33^***^ | [1.13,1.56] | 1.18^***^ | [1.08,1.30] |
| Fourth quartile (highest income) | 1.46^***^ | [1.22,1.74] | 1.39^***^ | [1.26,1.54] |
| *Household:* |  |  |  |  |
| Another person in household | 1.28^**^ | [1.10,1.49] | 1.34^***^ | [1.22,1.46] |
| Smoker in household | 0.90 | [0.78,1.03] | 0.88^**^ | [0.81,0.96] |
| Lives with spouse | 0.87 | [0.67,1.12] | 1.03 | [0.89,1.19] |
| Lives with child | 1.14 | [0.86,1.53] | 1.26^**^ | [1.07,1.48] |
| Lives with parent | 0.96 | [0.76,1.22] | 1.11 | [0.98,1.27] |
| Lives with sibling | 0.68^***^ | [0.59,0.79] | 0.71^***^ | [0.65,0.78] |
| Lives with other | 0.88 | [0.66,1.16] | 1.02 | [0.87,1.19] |
| Smoker in household – spouse | 0.86 | [0.63,1.19] | 0.70^***^ | [0.59,0.84] |
| Smoker in household – child | 1.01 | [0.70,1.47] | 0.83 | [0.67,1.03] |
| Smoker in household – parent | 0.84 | [0.59,1.19] | 0.80^*^ | [0.66,0.98] |
| Smoker in household – sibling | 1.28^**^ | [1.10,1.49] | 1.34^***^ | [1.22,1.46] |
| Smoker in household – other | 0.90 | [0.78,1.03] | 0.88^**^ | [0.81,0.96] |
| *Wave fixed effects:* |  |  |  |  |
| wave=3 (ref) | - | - | 1.00 | [1.00,1.00] |
| wave=4 | - | - | 0.88 | [0.74,1.05] |
| wave=5 | - | - | 0.88 | [0.74,1.05] |
| wave=6 | - | - | 1.08 | [0.91,1.28] |
| wave=7 | - | - | 0.84 | [0.70,1.00] |
| wave=8 | - | - | 0.89 | [0.74,1.06] |
| wave=9 | - | - | 0.98 | [0.82,1.17] |
| wave=10 | - | - | 1.00 | [0.84,1.20] |
| wave=11 | - | - | 1.05 | [0.88,1.26] |
| wave=12 | - | - | 1.12 | [0.94,1.32] |
| wave=13 | - | - | 1.14 | [0.96,1.35] |
| wave=14 | - | - | 0.99 | [0.83,1.18] |
| wave=15 | - | - | 0.96 | [0.81,1.14] |
| wave=16 | - | - | 0.85 | [0.71,1.02] |
| wave=17 | - | - | 1.00 | [0.84,1.18] |
| wave=18 | - | - | 0.94 | [0.79,1.12] |
| wave=19 | - | - | 0.93 | [0.78,1.11] |
| Constant | 0.10^***^ | [0.08,0.12] | 0.10^***^ | [0.09,0.12] |
| *N* | 11595.00 |  | 37096.00 |  |
| *Mean of outcome* | 0.15 |  | 0.14 |  |

Table A. 1 – Relapse models when restricting sample to 2014 to 2019 (1) and including year fixed effects (2)

|  | (1) |  | (2) |  |
| --- | --- | --- | --- | --- |
|  | **Relapse** |  | **Relapse** |  |
|  | $OR$ | [95% CI] | $OR$ | [95% CI] |
| *Consecutive waves abstained:* |  |  |  |  |
| 1-2 (ref) | 1.00 | [1.00,1.00] | 1.00 | [1.00,1.00] |
| 2-3 | 0.38^***^ | [0.31,0.46] | 0.37^***^ | [0.33,0.42] |
| 3-4 | 0.22^***^ | [0.18,0.28] | 0.22^***^ | [0.18,0.26] |
| 4-7 | 0.12^***^ | [0.10,0.15] | 0.10^***^ | [0.09,0.12] |
| 7 or more | 0.04^***^ | [0.03,0.05] | 0.04^***^ | [0.03,0.05] |
| *Level of smoking:* |  |  |  |  |
| 10 or fewer cigarettes per day (Ref) | 1.00 | [1.00,1.00] | 1.00 | [1.00,1.00] |
| 11-19 cigarettes per day | 1.13 | [0.93,1.37] | 1.15^*^ | [1.01,1.30] |
| 20 or more cigarettes per day | 1.15 | [0.91,1.45] | 1.06 | [0.91,1.23] |
| *Sex:* |  |  |  |  |
| Male | 1.03 | [0.89,1.19] | 1.09 | [0.99,1.20] |
| *Age (years):* |  |  |  |  |
| Under 30 | 1.15 | [0.90,1.47] | 1.14 | [0.97,1.33] |
| 30-39 | 1.17 | [0.92,1.48] | 1.05 | [0.90,1.23] |
| 40-49 | 1.15 | [0.89,1.49] | 1.07 | [0.91,1.25] |
| 50-59 (Ref) | 1.00 | [1.00,1.00] | 1.00 | [1.00,1.00] |
| 60-59 | 0.79 | [0.59,1.06] | 0.72^***^ | [0.60,0.87] |
| Over 70 | 0.45^***^ | [0.29,0.70] | 0.54^***^ | [0.41,0.73] |
| *Education:* |  |  |  |  |
| Less than high school | 1.00 | [1.00,1.00] | 1.00 | [1.00,1.00] |
| High school or equivalent | 0.89 | [0.74,1.06] | 0.94 | [0.84,1.06] |
| University degree | 0.72^**^ | [0.58,0.88] | 0.83^**^ | [0.72,0.95] |
| *Equivalised household income:* |  |  |  |  |
| First quartile (lowest income) | 1.00 | [1.00,1.00] | 1.00 | [1.00,1.00] |
| Second quartile | 0.90 | [0.73,1.11] | 0.87^*^ | [0.76,1.00] |
| Third quartile | 0.78^*^ | [0.63,0.95] | 0.78^***^ | [0.68,0.90] |
| Fourth quartile (highest income) | 0.74^*^ | [0.59,0.94] | 0.78^***^ | [0.68,0.90] |
| *Household:* |  |  |  |  |
| Another person in household | 0.78^**^ | [0.65,0.93] | 0.77^***^ | [0.68,0.87] |
| Smoker in household | 1.11 | [0.93,1.32] | 1.08 | [0.96,1.21] |
| Lives with spouse | 0.93 | [0.66,1.33] | 1.13 | [0.90,1.41] |
| Lives with child | 0.88 | [0.57,1.35] | 0.77 | [0.59,1.01] |
| Lives with parent | 1.03 | [0.75,1.43] | 1.09 | [0.88,1.35] |
| Lives with sibling | 1.40^**^ | [1.13,1.74] | 1.47^***^ | [1.27,1.69] |
| Lives with other | 1.15 | [0.75,1.76] | 1.20 | [0.94,1.54] |
| Smoker in household – spouse | 1.74^*^ | [1.07,2.82] | 1.39 | [1.00,1.92] |
| Smoker in household – child | 1.38 | [0.74,2.54] | 1.42 | [0.97,2.09] |
| Smoker in household – parent | 1.48 | [0.85,2.57] | 1.19 | [0.82,1.73] |
| Smoker in household – sibling | 0.78^**^ | [0.65,0.93] | 0.77^***^ | [0.68,0.87] |
| Smoker in household – other | 1.11 | [0.93,1.32] | 1.08 | [0.96,1.21] |
| *Wave fixed effects:* |  |  |  |  |
| wave=4 (ref) |  |  | 1.00 | [1.00,1.00] |
| wave=5 |  |  | 0.88 | [0.64,1.22] |
| wave=6 |  |  | 0.68^*^ | [0.49,0.93] |
| wave=7 |  |  | 0.73 | [0.54,1.00] |
| wave=8 |  |  | 0.71^*^ | [0.51,0.98] |
| wave=9 |  |  | 0.65^**^ | [0.47,0.89] |
| wave=10 |  |  | 0.73^*^ | [0.53,0.99] |
| wave=11 |  |  | 0.65^**^ | [0.47,0.88] |
| wave=12 |  |  | 0.66^**^ | [0.49,0.90] |
| wave=13 |  |  | 0.63^**^ | [0.46,0.85] |
| wave=14 |  |  | 0.70^*^ | [0.52,0.94] |
| wave=15 |  |  | 0.70^*^ | [0.51,0.94] |
| wave=16 |  |  | 0.77 | [0.58,1.04] |
| wave=17 |  |  | 0.71^*^ | [0.52,0.95] |
| wave=18 |  |  | 0.80 | [0.60,1.08] |
| wave=19 |  |  | 0.76 | [0.56,1.02] |
| Constant | 0.72^*^ | [0.55,0.96] | 0.96 | [0.71,1.29] |
| *N* | *8,443* |  | *17,478* |  |
| *Mean of outcome* | *0.12* |  | *0.15* |  |

#### SM.5. Classifying individuals who smoked less than weekly or smoking weekly but less than daily as non-smokers

|  | (1) |  | (2) |  | (3) |  |
| --- | --- | --- | --- | --- | --- | --- |
|  | **Quit** |  | **Quit** |  | **Quit** |  |
|  | $OR$ | [95% CI] | $OR$ | [95% CI] | $OR$ | [95% CI] |
| Previous quit recorded | 2.00^***^ | [1.85,2.16] | 1.99^***^ | [1.84,2.15] | 2.02^***^ | [1.87,2.19] |
| *Level of smoking:* |  |  |  |  |  |  |
| 10 or fewer cigarettes per day (Ref) | 1.00 | [1.00,1.00] | 1.00 | [1.00,1.00] | 1.00 | [1.00,1.00] |
| 11-19 cigarettes per day | 0.61^***^ | [0.56,0.66] | 0.61^***^ | [0.56,0.66] | 0.61^***^ | [0.56,0.66] |
| 20 or more cigarettes per day | 0.50^***^ | [0.45,0.55] | 0.50^***^ | [0.46,0.55] | 0.51^***^ | [0.46,0.56] |
| *Sex:* |  |  |  |  |  |  |
| Male | 1.00 | [0.93,1.08] | 1.00 | [0.93,1.08] | 0.96 | [0.89,1.04] |
| *Age (years):* |  |  |  |  |  |  |
| Under 30 | 1.58^***^ | [1.41,1.76] | 1.59^***^ | [1.42,1.78] | 1.62^***^ | [1.44,1.83] |
| 30-39 | 1.22^***^ | [1.09,1.37] | 1.22^***^ | [1.09,1.37] | 1.31^***^ | [1.16,1.47] |
| 40-49 | 0.99 | [0.88,1.11] | 0.99 | [0.88,1.11] | 1.03 | [0.92,1.16] |
| 50-59 (Ref) | 1.00 | [1.00,1.00] | 1.00 | [1.00,1.00] | 1.00 | [1.00,1.00] |
| 60-59 | 1.32^***^ | [1.14,1.52] | 1.31^***^ | [1.14,1.51] | 1.27^**^ | [1.10,1.46] |
| Over 70 | 1.33^*^ | [1.07,1.65] | 1.31^*^ | [1.05,1.63] | 1.25^*^ | [1.01,1.55] |
| *Education:* |  |  |  |  |  |  |
| Less than high school | 1.00 | [1.00,1.00] | 1.00 | [1.00,1.00] | 1.00 | [1.00,1.00] |
| High school or equivalent | 1.13^**^ | [1.04,1.22] | 1.13^**^ | [1.04,1.22] | 1.13^**^ | [1.05,1.23] |
| University degree | 1.34^***^ | [1.20,1.49] | 1.34^***^ | [1.20,1.49] | 1.35^***^ | [1.21,1.50] |
| *Equivalised household income:* |  |  |  |  |  |  |
| First quartile (lowest income) | 1.00 | [1.00,1.00] | 1.00 | [1.00,1.00] | 1.00 | [1.00,1.00] |
| Second quartile | 0.97 | [0.88,1.06] | 0.96 | [0.88,1.05] | 0.96 | [0.87,1.05] |
| Third quartile | 1.15^**^ | [1.04,1.26] | 1.14^**^ | [1.03,1.25] | 1.10 | [1.00,1.21] |
| Fourth quartile (highest income) | 1.37^***^ | [1.23,1.51] | 1.34^***^ | [1.21,1.49] | 1.27^***^ | [1.15,1.42] |
| *Household:* |  |  |  |  |  |  |
| Another person in household |  |  | 1.15^**^ | [1.04,1.28] |  |  |
| Smoker in household |  |  | 0.84^***^ | [0.78,0.91] |  |  |
| Lives with spouse |  |  |  |  | 1.27^***^ | [1.16,1.40] |
| Lives with child |  |  |  |  | 0.87^**^ | [0.80,0.94] |
| Lives with parent |  |  |  |  | 1.00 | [0.86,1.17] |
| Lives with sibling |  |  |  |  | 1.30^**^ | [1.09,1.55] |
| Lives with other |  |  |  |  | 1.13 | [0.98,1.30] |
| Smoker in household – spouse |  |  |  |  | 0.76^***^ | [0.70,0.84] |
| Smoker in household – child |  |  |  |  | 1.18 | [0.99,1.39] |
| Smoker in household – parent |  |  |  |  | 0.83 | [0.69,1.01] |
| Smoker in household – sibling |  |  |  |  | 0.80 | [0.61,1.03] |
| Smoker in household – other |  |  |  |  | 0.78^*^ | [0.63,0.97] |
| Constant | 0.13^***^ | [0.11,0.15] | 0.12^***^ | [0.11,0.14] | 0.13^***^ | [0.11,0.15] |
| *N* | *30,466* |  | *30,466* |  | *30,466* |  |
| *Mean of outcome* | *0.15* |  | *0.15* |  | *0.15* |  |
|  | (1) |  | (2) |  | (3) |  |
|  | **Relapse** |  | **Relapse** |  | **Relapse** |  |
|  | $OR$ | [95% CI] | $OR$ | [95% CI] | $OR$ | [95% CI] |
| *Consecutive waves abstained:* |  |  |  |  |  |  |
| 1-2 (ref) | 1.00 | [1.00,1.00] | 1.00 | [1.00,1.00] | 1.00 | [1.00,1.00] |
| 2-3 | 0.35^***^ | [0.31,0.40] | 0.35^***^ | [0.31,0.40] | 0.36^***^ | [0.32,0.40] |
| 3-4 | 0.19^***^ | [0.17,0.23] | 0.20^***^ | [0.17,0.23] | 0.20^***^ | [0.17,0.24] |
| 4-7 | 0.09^***^ | [0.08,0.11] | 0.09^***^ | [0.08,0.11] | 0.09^***^ | [0.08,0.11] |
| 7 or more | 0.03^***^ | [0.02,0.04] | 0.03^***^ | [0.03,0.04] | 0.03^***^ | [0.03,0.04] |
| *Level of smoking:* |  |  |  |  |  |  |
| 10 or fewer cigarettes per day (Ref) | 1.00 | [1.00,1.00] | 1.00 | [1.00,1.00] | 1.00 | [1.00,1.00] |
| 11-19 cigarettes per day | 0.86^*^ | [0.76,0.98] | 0.87^*^ | [0.77,0.99] | 0.87^*^ | [0.77,0.99] |
| 20 or more cigarettes per day | 0.88 | [0.76,1.01] | 0.87 | [0.75,1.01] | 0.88 | [0.76,1.02] |
| *Sex:* |  |  |  |  |  |  |
| Male | 0.99 | [0.90,1.08] | 1.00 | [0.91,1.10] | 1.03 | [0.94,1.14] |
| *Age (years):* |  |  |  |  |  |  |
| Under 30 | 1.18^*^ | [1.02,1.37] | 1.17^*^ | [1.00,1.36] | 1.12 | [0.95,1.31] |
| 30-39 | 0.95 | [0.81,1.10] | 0.96 | [0.83,1.12] | 0.94 | [0.81,1.11] |
| 40-49 | 1.08 | [0.93,1.26] | 1.09 | [0.93,1.26] | 1.06 | [0.91,1.24] |
| 50-59 (Ref) | 1.00 | [1.00,1.00] | 1.00 | [1.00,1.00] | 1.00 | [1.00,1.00] |
| 60-59 | 0.71^***^ | [0.59,0.86] | 0.73^**^ | [0.60,0.88] | 0.74^**^ | [0.61,0.90] |
| Over 70 | 0.55^***^ | [0.42,0.71] | 0.57^***^ | [0.43,0.74] | 0.59^***^ | [0.45,0.76] |
| *Education:* |  |  |  |  |  |  |
| Less than high school | 1.00 | [1.00,1.00] | 1.00 | [1.00,1.00] | 1.00 | [1.00,1.00] |
| High school or equivalent | 0.90 | [0.81,1.01] | 0.92 | [0.82,1.03] | 0.91 | [0.82,1.02] |
| University degree | 0.74^***^ | [0.64,0.84] | 0.76^***^ | [0.67,0.87] | 0.76^***^ | [0.66,0.87] |
| *Equivalised household income:* |  |  |  |  |  |  |
| First quartile (lowest income) | 1.00 | [1.00,1.00] | 1.00 | [1.00,1.00] | 1.00 | [1.00,1.00] |
| Second quartile | 0.82^**^ | [0.72,0.94] | 0.82^**^ | [0.72,0.93] | 0.83^**^ | [0.73,0.95] |
| Third quartile | 0.74^***^ | [0.65,0.84] | 0.73^***^ | [0.64,0.83] | 0.76^***^ | [0.67,0.87] |
| Fourth quartile (highest income) | 0.64^***^ | [0.55,0.73] | 0.64^***^ | [0.55,0.74] | 0.69^***^ | [0.59,0.80] |
| *Household:* |  |  |  |  |  |  |
| Another person in household |  |  | 0.93 | [0.82,1.06] |  |  |
| Smoker in household |  |  | 1.51^***^ | [1.34,1.69] |  |  |
| Lives with spouse |  |  |  |  | 0.76^***^ | [0.67,0.85] |
| Lives with child |  |  |  |  | 1.08 | [0.96,1.22] |
| Lives with parent |  |  |  |  | 1.13 | [0.90,1.42] |
| Lives with sibling |  |  |  |  | 0.82 | [0.62,1.08] |
| Lives with other |  |  |  |  | 1.10 | [0.89,1.36] |
| Smoker in household – spouse |  |  |  |  | 1.66^***^ | [1.44,1.91] |
| Smoker in household – child |  |  |  |  | 1.21 | [0.93,1.59] |
| Smoker in household – parent |  |  |  |  | 1.38 | [0.98,1.94] |
| Smoker in household – sibling |  |  |  |  | 1.41 | [0.96,2.07] |
| Smoker in household – other |  |  |  |  | 1.01 | [0.69,1.50] |
| Constant | 0.97 | [0.82,1.14] | 0.91 | [0.76,1.09] | 0.94 | [0.79,1.12] |
| *N* | 16356.00 |  | 16356.00 |  | 16356.00 |  |
| *Mean of outcome* | 0.16 |  | 0.16 |  | 0.16 |  |

#### SM.6. Classifying a quit attempt as two waves abstained from smoking

|  | (1) |  | (2) |  | (3) |  |
| --- | --- | --- | --- | --- | --- | --- |
|  | **Quit** |  | **Quit** |  | **Quit** |  |
|  | $OR$ | [95% CI] | $OR$ | [95% CI] | $OR$ | [95% CI] |
| Previous quit recorded | 6.28^***^ | [5.21,7.58] | 6.06^***^ | [5.02,7.32] | 6.31^***^ | [5.21,7.64] |
| *Level of smoking:* |  |  |  |  |  |  |
| 10 or fewer cigarettes per day (Ref) | 1.00 | [1.00,1.00] | 1.00 | [1.00,1.00] | 1.00 | [1.00,1.00] |
| 11-19 cigarettes per day | 0.37^***^ | [0.29,0.47] | 0.38^***^ | [0.30,0.49] | 0.39^***^ | [0.31,0.50] |
| 20 or more cigarettes per day | 0.33^***^ | [0.26,0.43] | 0.36^***^ | [0.28,0.47] | 0.38^***^ | [0.29,0.49] |
| *Sex:* |  |  |  |  |  |  |
| Male | 1.09 | [0.90,1.31] | 1.07 | [0.88,1.29] | 0.98 | [0.81,1.19] |
| *Age (years):* |  |  |  |  |  |  |
| Under 30 | 1.00 | [1.00,1.00] | 1.00 | [1.00,1.00] | 1.00 | [1.00,1.00] |
| 30-39 | 1.03 | [0.81,1.30] | 0.96 | [0.76,1.21] | 0.87 | [0.67,1.13] |
| 40-49 | 0.78 | [0.60,1.01] | 0.70^**^ | [0.54,0.92] | 0.61^***^ | [0.46,0.81] |
| 50-59 (Ref) | 0.80 | [0.60,1.06] | 0.74^*^ | [0.55,0.98] | 0.59^***^ | [0.43,0.80] |
| 60-59 | 1.01 | [0.72,1.42] | 0.89 | [0.63,1.25] | 0.67^*^ | [0.47,0.96] |
| Over 70 | 0.67 | [0.36,1.24] | 0.54^*^ | [0.29,0.99] | 0.39^**^ | [0.21,0.72] |
| *Education:* |  |  |  |  |  |  |
| Less than high school | 1.00 | [1.00,1.00] | 1.00 | [1.00,1.00] | 1.00 | [1.00,1.00] |
| High school or equivalent | 1.32^*^ | [1.06,1.65] | 1.28^*^ | [1.02,1.60] | 1.29^*^ | [1.02,1.62] |
| University degree | 1.85^***^ | [1.43,2.38] | 1.76^***^ | [1.36,2.27] | 1.79^***^ | [1.38,2.32] |
| *Equivalised household income:* |  |  |  |  |  |  |
| First quartile (lowest income) | 1.00 | [1.00,1.00] | 1.00 | [1.00,1.00] | 1.00 | [1.00,1.00] |
| Second quartile | 0.90 | [0.71,1.14] | 0.90 | [0.71,1.15] | 0.86 | [0.67,1.09] |
| Third quartile | 1.05 | [0.83,1.33] | 1.04 | [0.82,1.32] | 0.95 | [0.75,1.21] |
| Fourth quartile (highest income) | 1.26 | [0.98,1.61] | 1.25 | [0.97,1.61] | 1.10 | [0.85,1.43] |
| *Household:* |  |  |  |  |  |  |
| Another person in household |  |  | 1.65^***^ | [1.31,2.10] |  |  |
| Smoker in household |  |  | 0.36^***^ | [0.30,0.45] |  |  |
| Lives with spouse |  |  |  |  | 2.15^***^ | [1.72,2.67] |
| Lives with child |  |  |  |  | 0.90 | [0.73,1.11] |
| Lives with parent |  |  |  |  | 0.91 | [0.59,1.40] |
| Lives with sibling |  |  |  |  | 1.00 | [0.60,1.66] |
| Lives with other |  |  |  |  | 1.61^**^ | [1.15,2.26] |
| Smoker in household – spouse |  |  |  |  | 0.27^***^ | [0.21,0.35] |
| Smoker in household – child |  |  |  |  | 0.91 | [0.61,1.37] |
| Smoker in household – parent |  |  |  |  | 0.34^**^ | [0.17,0.69] |
| Smoker in household – sibling |  |  |  |  | 1.11 | [0.58,2.13] |
| Smoker in household – other |  |  |  |  | 0.31^**^ | [0.15,0.62] |
| Constant | 0.02^***^ | [0.02,0.03] | 0.02^***^ | [0.02,0.03] | 0.03^***^ | [0.02,0.04] |
| *N* | *24,239* |  | *24,239* |  | *24,239* |  |
| *Mean of outcome* | *0.03* |  | *0.03* |  | *0.03* |  |

#### SM.7. Disaggregating household members by under 15 years

Starred columns present disaggregation by whether the child or sibling is under 15 years old.

|  | (2) |  | (2*) |  |
| --- | --- | --- | --- | --- |
|  | **Quit** |  | **Quit** |  |
|  | $OR$ | [95% CI] | $OR$ | [95% CI] |
| Previous quit recorded | 2.22^***^ | [2.06,2.39] | 2.22^***^ | [2.06,2.40] |
| *Level of smoking:* |  |  |  |  |
| 10 or fewer cigarettes per day (Ref) | 1.00 | [1.00,1.00] | 1.00 | [1.00,1.00] |
| 11-19 cigarettes per day | 0.55^***^ | [0.51,0.60] | 0.55^***^ | [0.51,0.60] |
| 20 or more cigarettes per day | 0.47^***^ | [0.43,0.52] | 0.47^***^ | [0.43,0.52] |
| *Sex:* |  |  |  |  |
| Male | 0.99 | [0.92,1.06] | 0.99 | [0.92,1.06] |
| *Age (years):* |  |  |  |  |
| Under 30 | 1.62^***^ | [1.45,1.80] | 1.62^***^ | [1.46,1.80] |
| 30-39 | 1.25^***^ | [1.12,1.39] | 1.26^***^ | [1.13,1.41] |
| 40-49 | 0.98 | [0.88,1.09] | 0.98 | [0.88,1.10] |
| 50-59 (Ref) | 1.00 | [1.00,1.00] | 1.00 | [1.00,1.00] |
| 60-59 | 1.31^***^ | [1.14,1.51] | 1.31^***^ | [1.14,1.50] |
| Over 70 | 1.29^*^ | [1.04,1.59] | 1.28^*^ | [1.04,1.59] |
| *Education:* |  |  |  |  |
| Less than high school | 1.00 | [1.00,1.00] | 1.00 | [1.00,1.00] |
| High school or equivalent | 1.14^**^ | [1.05,1.23] | 1.14^**^ | [1.05,1.23] |
| University degree | 1.42^***^ | [1.29,1.56] | 1.42^***^ | [1.29,1.56] |
| *Equivalised household income:* |  |  |  |  |
| First quartile (lowest income) | 1.00 | [1.00,1.00] | 1.00 | [1.00,1.00] |
| Second quartile | 1.06 | [0.97,1.16] | 1.06 | [0.97,1.16] |
| Third quartile | 1.23^***^ | [1.12,1.35] | 1.22^***^ | [1.11,1.34] |
| Fourth quartile (highest income) | 1.47^***^ | [1.34,1.62] | 1.46^***^ | [1.32,1.62] |
| *Household:* |  |  |  |  |
| Another person in household | 1.22^***^ | [1.11,1.34] | 1.23^***^ | [1.12,1.35] |
| Another person in household (under 15) | - | - | 0.98 | [0.90,1.06] |
| Smoker in household | 0.77^***^ | [0.72,0.83] | 0.77^***^ | [0.72,0.83] |
| Constant | 0.10^***^ | [0.08,0.11] | 0.10^***^ | [0.08,0.11] |
| *N* | *37,096* |  | *37,096* |  |
| *Mean of outcome* | *0.14* |  | *0.14* |  |

|  | (3) |  | (3*) |  |
| --- | --- | --- | --- | --- |
|  | **Quit** |  | **Quit** |  |
|  | $OR$ | [95% CI] | $OR$ | [95% CI] |
| Previous quit recorded | 2.24^***^ | [2.08,2.42] | 2.24^***^ | [2.08,2.42] |
| *Level of smoking:* |  |  |  |  |
| 10 or fewer cigarettes per day (Ref) | 1.00 | [1.00,1.00] | 1.00 | [1.00,1.00] |
| 11-19 cigarettes per day | 0.55^***^ | [0.51,0.60] | 0.55^***^ | [0.51,0.60] |
| 20 or more cigarettes per day | 0.48^***^ | [0.43,0.53] | 0.48^***^ | [0.43,0.53] |
| *Sex:* |  |  |  |  |
| Male | 0.95 | [0.89,1.02] | 0.95 | [0.89,1.02] |
| *Age (years):* |  |  |  |  |
| Under 30 | 1.66^***^ | [1.48,1.86] | 1.60^***^ | [1.42,1.80] |
| 30-39 | 1.32^***^ | [1.18,1.48] | 1.27^***^ | [1.13,1.42] |
| 40-49 | 1.02 | [0.92,1.14] | 1.02 | [0.91,1.13] |
| 50-59 (Ref) | 1.00 | [1.00,1.00] | 1.00 | [1.00,1.00] |
| 60-59 | 1.27^***^ | [1.10,1.45] | 1.25^**^ | [1.09,1.44] |
| Over 70 | 1.22 | [0.99,1.51] | 1.20 | [0.97,1.48] |
| *Education:* |  |  |  |  |
| Less than high school | 1.00 | [1.00,1.00] | 1.00 | [1.00,1.00] |
| High school or equivalent | 1.14^**^ | [1.05,1.23] | 1.14^**^ | [1.05,1.23] |
| University degree | 1.41^***^ | [1.28,1.56] | 1.41^***^ | [1.28,1.55] |
| *Equivalised household income:* |  |  |  |  |
| First quartile (lowest income) | 1.00 | [1.00,1.00] | 1.00 | [1.00,1.00] |
| Second quartile | 1.05 | [0.96,1.15] | 1.05 | [0.96,1.15] |
| Third quartile | 1.18^***^ | [1.08,1.30] | 1.19^***^ | [1.08,1.30] |
| Fourth quartile (highest income) | 1.39^***^ | [1.26,1.53] | 1.40^***^ | [1.27,1.54] |
| *Household:* |  |  |  |  |
| Lives with spouse | 1.34^***^ | [1.23,1.46] | 1.34^***^ | [1.22,1.46] |
| Lives with child | 0.88^**^ | [0.81,0.96] | 0.93 | [0.85,1.02] |
| Lives with child (over 15) | - | - | 0.84^**^ | [0.74,0.95] |
| Lives with parent | 1.02 | [0.89,1.18] | 1.03 | [0.89,1.19] |
| Lives with sibling | 1.26^**^ | [1.07,1.48] | 1.12 | [0.86,1.46] |
| Lives with sibling (over 15) | - | - | 1.17 | [0.89,1.55] |
| Lives with other | 1.12 | [0.98,1.27] | 0.96 | [0.76,1.22] |
| Lives with other (over 15) | - | - | 1.25 | [0.95,1.65] |
| Smoker in household – spouse | 0.71^***^ | [0.65,0.78] | 0.71^***^ | [0.65,0.78] |
| Smoker in household – child | 1.02 | [0.87,1.19] | 1.14 | [0.96,1.37] |
| Smoker in household – parent | 0.71^***^ | [0.59,0.84] | 0.71^***^ | [0.59,0.85] |
| Smoker in household – sibling | 0.83 | [0.67,1.03] | 0.80 | [0.64,1.01] |
| Smoker in household – other | 0.80^*^ | [0.66,0.98] | 0.75^*^ | [0.60,0.93] |
| Constant | 0.10^***^ | [0.09,0.11] | 0.10^***^ | [0.09,0.12] |
| *N* | *37,096* |  | *37,096* |  |
| *Mean of outcome* | *0.14* |  | *0.14* |  |

|  | (2) |  | (2*) |  |
| --- | --- | --- | --- | --- |
|  | **Relapse** |  | **Relapse** |  |
|  | $OR$ | [95% CI] | $OR$ | [95% CI] |
| *Consecutive waves abstained:* |  |  |  |  |
| 1-2 (ref) | 1.00 | [1.00,1.00] | 1.00 | [1.00,1.00] |
| 2-3 | 0.36^***^ | [0.32,0.41] | 0.36^***^ | [0.32,0.41] |
| 3-4 | 0.21^***^ | [0.18,0.25] | 0.21^***^ | [0.18,0.25] |
| 4-7 | 0.10^***^ | [0.09,0.12] | 0.10^***^ | [0.09,0.12] |
| 7 or more | 0.04^***^ | [0.03,0.05] | 0.04^***^ | [0.03,0.05] |
| *Level of smoking:* |  |  |  |  |
| 10 or fewer cigarettes per day (Ref) | 1.00 | [1.00,1.00] | 1.00 | [1.00,1.00] |
| 11-19 cigarettes per day | 1.14^*^ | [1.00,1.29] | 1.14^*^ | [1.00,1.29] |
| 20 or more cigarettes per day | 1.05 | [0.91,1.22] | 1.06 | [0.91,1.22] |
| *Sex:* |  |  |  |  |
| Male | 1.06 | [0.97,1.16] | 1.06 | [0.96,1.16] |
| *Age (years):* |  |  |  |  |
| Under 30 | 1.18^*^ | [1.02,1.37] | 1.19^*^ | [1.03,1.38] |
| 30-39 | 1.06 | [0.91,1.23] | 1.09 | [0.93,1.27] |
| 40-49 | 1.08 | [0.93,1.25] | 1.10 | [0.94,1.29] |
| 50-59 (Ref) | 1.00 | [1.00,1.00] | 1.00 | [1.00,1.00] |
| 60-59 | 0.71^***^ | [0.59,0.85] | 0.70^***^ | [0.58,0.85] |
| Over 70 | 0.53^***^ | [0.40,0.71] | 0.53^***^ | [0.40,0.70] |
| *Education:* |  |  |  |  |
| Less than high school | 1.00 | [1.00,1.00] | 1.00 | [1.00,1.00] |
| High school or equivalent | 0.95 | [0.84,1.06] | 0.95 | [0.84,1.06] |
| University degree | 0.83^**^ | [0.72,0.95] | 0.83^**^ | [0.72,0.95] |
| *Equivalised household income:* |  |  |  |  |
| First quartile (lowest income) | 1.00 | [1.00,1.00] | 1.00 | [1.00,1.00] |
| Second quartile | 0.84^*^ | [0.73,0.96] | 0.84^*^ | [0.74,0.96] |
| Third quartile | 0.74^***^ | [0.65,0.85] | 0.74^***^ | [0.65,0.85] |
| Fourth quartile (highest income) | 0.72^***^ | [0.63,0.83] | 0.71^***^ | [0.62,0.82] |
| *Household:* |  |  |  |  |
| Another person in household | 1.01 | [0.89,1.15] | 1.03 | [0.90,1.18] |
| Another person in household (under 15) | - | - | 0.95 | [0.84,1.07] |
| Smoker in household | 1.37^***^ | [1.22,1.53] | 1.37^***^ | [1.22,1.53] |
| Constant | 0.64^***^ | [0.53,0.78] | 0.64^***^ | [0.53,0.78] |
| *N* | *17,478* |  | *17,478* |  |
| *Mean of outcome* | *0.15* |  | *0.15* |  |

|  | (3) |  | (3*) |  |
| --- | --- | --- | --- | --- |
|  | **Relapse** |  | **Relapse** |  |
|  | $OR$ | [95% CI] | $OR$ | [95% CI] |
| *Consecutive waves abstained:* |  |  |  |  |
| 1-2 (ref) | 1.00 | [1.00,1.00] | 1.00 | [1.00,1.00] |
| 2-3 | 0.36^***^ | [0.32,0.41] | 0.36*** | [0.32,0.41] |
| 3-4 | 0.21^***^ | [0.18,0.25] | 0.21*** | [0.18,0.25] |
| 4-7 | 0.10^***^ | [0.09,0.12] | 0.10*** | [0.09,0.12] |
| 7 or more | 0.04^***^ | [0.03,0.05] | 0.04*** | [0.03,0.05] |
| *Level of smoking:* |  |  |  |  |
| 10 or fewer cigarettes per day (Ref) | 1.00 | [1.00,1.00] | 1.00 | [1.00,1.00] |
| 11-19 cigarettes per day | 1.14^*^ | [1.00,1.29] | 1.14^*^ | [1.00,1.29] |
| 20 or more cigarettes per day | 1.05 | [0.91,1.22] | 1.06 | [0.91,1.22] |
| *Sex:* |  |  |  |  |
| Male | 1.09 | [0.99,1.20] | 1.09 | [0.99,1.20] |
| *Age (years):* |  |  |  |  |
| Under 30 | 1.14 | [0.97,1.33] | 1.14 | [0.97,1.34] |
| 30-39 | 1.06 | [0.91,1.24] | 1.06 | [0.90,1.24] |
| 40-49 | 1.07 | [0.91,1.25] | 1.07 | [0.91,1.25] |
| 50-59 (Ref) | 1.00 | [1.00,1.00] | 1.00 | [1.00,1.00] |
| 60-59 | 0.72^***^ | [0.60,0.87] | 0.72^***^ | [0.60,0.87] |
| Over 70 | 0.55^***^ | [0.41,0.73] | 0.55^***^ | [0.41,0.73] |
| *Education:* |  |  |  |  |
| Less than high school | 1.00 | [1.00,1.00] | 1.00 | [1.00,1.00] |
| High school or equivalent | 0.95 | [0.84,1.06] | 0.95 | [0.84,1.06] |
| University degree | 0.82^**^ | [0.72,0.94] | 0.82^**^ | [0.72,0.94] |
| *Equivalised household income:* |  |  |  |  |
| First quartile (lowest income) | 1.00 | [1.00,1.00] | 1.00 | [1.00,1.00] |
| Second quartile | 0.86^*^ | [0.75,0.99] | 0.86^*^ | [0.75,0.99] |
| Third quartile | 0.78^***^ | [0.68,0.89] | 0.78^***^ | [0.68,0.89] |
| Fourth quartile (highest income) | 0.78^***^ | [0.67,0.90] | 0.78^***^ | [0.67,0.90] |
| *Household:* |  |  |  |  |
| Lives with spouse | 0.77^***^ | [0.68,0.87] | 0.77^***^ | [0.68,0.87] |
| Lives with child | 1.08 | [0.96,1.21] | 1.08 | [0.95,1.22] |
| Lives with child (over 15) | - | - | 1.00 | [0.84,1.19] |
| Lives with parent | 1.12 | [0.90,1.40] | 1.12 | [0.89,1.40] |
| Lives with sibling | 0.77 | [0.59,1.01] | 0.78 | [0.48,1.27] |
| Lives with sibling (over 15) | - | - | 0.98 | [0.59,1.64] |
| Lives with other | 1.08 | [0.87,1.34] | 1.22 | [0.83,1.81] |
| Lives with other (over 15) | - | - | 0.85 | [0.54,1.33] |
| Smoker in household – spouse | 1.47^***^ | [1.28,1.69] | 1.47^***^ | [1.28,1.69] |
| Smoker in household – child | 1.20 | [0.94,1.53] | 1.19 | [0.91,1.55] |
| Smoker in household – parent | 1.39^*^ | [1.00,1.94] | 1.40^*^ | [1.00,1.94] |
| Smoker in household – sibling | 1.43 | [0.97,2.11] | 1.43 | [0.96,2.13] |
| Smoker in household – other | 1.20 | [0.82,1.74] | 1.25 | [0.84,1.86] |
| Constant | 0.70^***^ | [0.59,0.84] | 0.70^***^ | [0.59,0.84] |
| *N* | *17,478* |  | *17,478* |  |
| *Mean of outcome* | *0.15* |  | *0.15* |  |
